# Interregional SARS-CoV-2 spread from a single introduction outbreak in a meat-packing plant in northeast Iowa

**DOI:** 10.1101/2020.06.08.20125534

**Authors:** Craig S. Richmond, Arick P. Sabin, Dean A. Jobe, Steven D. Lovrich, Paraic A. Kenny

## Abstract

SARS-CoV-2 spread has proven to be especially difficult to mitigate in high risk settings, including nursing homes, cruises, prisons and various industrial settings. Among industrial settings, meat processing facilities in the United States have experienced particularly challenging outbreaks. We have sequenced SARS-CoV-2 whole viral genomes from individuals testing positive in an integrated regional healthcare system serving 21 counties in southwestern Wisconsin, northeastern Iowa and southeastern Minnesota, providing an overview of SARS-CoV-2 introduction and spread in a region spanning multiple jurisdictions with differing mitigation policies. While most viral introductions we detected were contained with only minor transmission chains, a striking exception was an outbreak associated with a meatpacking plant in Postville, IA. In this case, a single viral introduction led to unrestrained spread within the facility, affecting many staff and members of their households. Importantly, by surveilling viral sequences from the surrounding counties, we have documented the spread of this SARS-CoV-2 substrain from this epicenter to individuals in 13 cities in 7 counties in Iowa, Wisconsin and Minnesota, a region spanning 185 square miles. This study highlights the regional public health consequences of failures to rapidly act to mitigate viral spread in a single industrial setting.

## INTRODUCTION

The SARS-CoV-2 betacoronavirus is a novel respiratory pathogen which emerged in Wuhan, China in late 2019 (1). An early assessment of the 29,903 base RNA genome sequence of 95 viral specimens highlighted several sequence variants (2). Numerous teams around the world have produced a rapidly increasing number of publicly available sequences (32,673 in GISAID on 5/27/20) with obvious sequence diversification as viral replication and spread has continued. These sequence variants, inherited in each cycle of viral replication, allow the generation of phylogenetic trees through which the evolution of the virus can be traced. Individual viruses acquire mutations during replication at an approximate rate of 1-2 sites per month (3). This mutation rate provides good resolution for both global and local tracking of viral substrains. Because viral replication occurs in individuals, tracking these inherited variants acts as a proxy for tracking the spread of the virus through populations of individuals.

In the present study, we have monitored the introduction and spread of SARS-CoV-2 to the service area of the Gundersen Health System, an integrated healthcare system headquartered in La Crosse, WI and providing care in 21 counties in southwestern Wisconsin, northeastern Iowa and southeastern Minnesota. We have performed whole viral genome sequencing on 67 positive cases from this region, spanning three states which have adopted mitigation approaches of differing intensity.

## MATERIALS AND METHODS

### Specimens

Specimens were nasopharyngeal swabs taken for diagnostic purposes and evaluated for SARS-CoV-2 positivity at the Gundersen Medical Foundation’s Molecular Diagnostics Laboratory using the CDC 2019-nCoV Real-Time RT-PCR Diagnostic assay. This study includes positive cases diagnosed between 3/18/2020 and 5/24/2020) Ethical approval was obtained from the Gundersen Health System Institutional Review Board (#2-20-03-008; PI: Kenny) to perform additional next-generation sequencing on remnant specimens after completion of diagnostic testing. Samples for this study include only those testing positive in Gundersen Healthcare System’s diagnostic laboratories and, as such, do not include other cases from the region which may have been tested in other healthcare systems and/or public health laboratories.

### RNA isolation and cDNA synthesis

Nasopharyngeal swabs were immersed in liquid medium. RNA was isolated using QIAmp Viral RNA Mini kit (Qiagen, Germantown, MD). cDNA was synthesized from 5 μl of purified RNA using ProtoScript II First Strand cDNA Synthesis Kit (New England Biolabs, Ipswich, MA). Early libraries were generated from cDNA prepared using a mixture of random hexamers and oligo dT, although we soon determined that omission of oligo dT led to better depth of coverage across the SARS-CoV-2 genome.

### Next Generation sequencing

We used the Ion AmpliSeq SARS-CoV-2 Panel (Thermo-Fisher, Waltham, MA) to sequence 237 viral specific targets encompassing the complete viral genome from cDNA derived from SARS-COV-2-positive clinical specimens. Barcoded multiplexed libraries were prepared in batches of eight on the Ion Chef (Thermo-Fisher) and sequenced on the Ion Torrent S5 (Thermo-Fisher), most commonly with a single library of 8 specimens on an Ion 520 chip. For samples with low viral load (typically CT values above 30), the merging of data from replicate runs was often required to achieve good coverage.

### Bioinformatics

Demultiplexed Tmap-aligned bam files were produced by the Ion Torrent S5. To overcome difficulties caused by read soft-clipping by this non-splice aware aligner, individual sequence reads were extracted using samtools (4) and re-aligned to the SARS-CoV-2 genome using a splice-aware aligner, hisat2 (5).

These bam files were subjected to QC review in IGV. Samples passing QC had a consensus sequence computed by IGV according to the method of Cavener (6). Consensus sequences were reported to GISAID (7). For phylogenetic inference (i.e. to determine the hierarchy of case relationships) samples were integrated with associated metadata and aligned on a local implementation of NextStrain (8) using augur and displayed via a web browser using auspice.

## RESULTS

Viral genome sequencing was attempted on all samples testing positive in Gundersen Healthcare System’s diagnostic laboratories. Good genome-wide coverage was typically achieved with specimens with clinical test Ct values up to 30/31, and merging of data from multiple runs allowed analysis of samples with higher Ct values. Consensus viral sequences were loaded into a local implementation of NextStrain, along with a background dataset of viral sequences from elsewhere in Wisconsin, Minnesota, Iowa and Illinois which was downloaded from GISAID (See Supplemental Table 1 for all sequences used in this study). Phylogenetic analysis indicated 14 independent viral introductions to our region (Fig 1).

**Figure 1.**
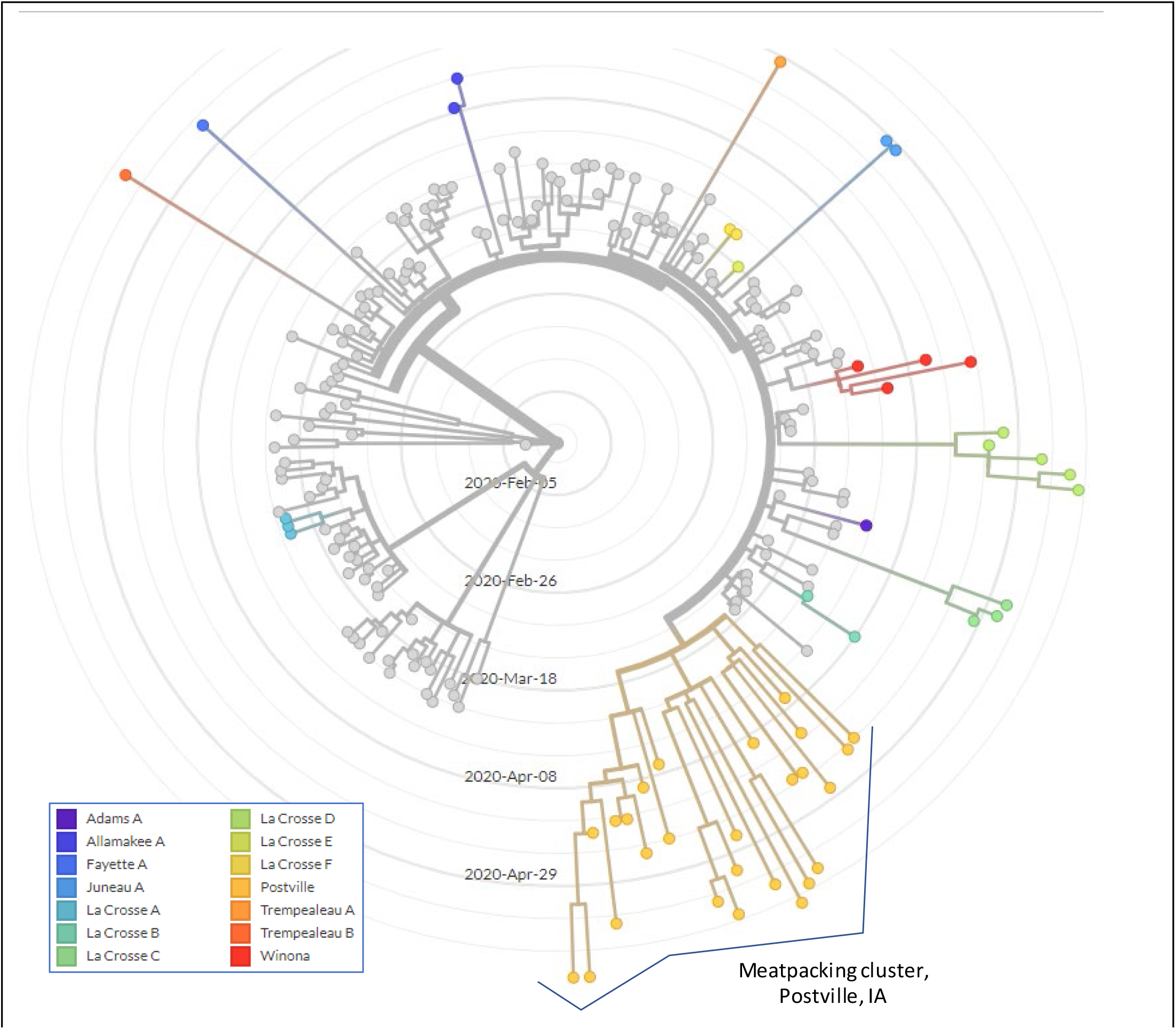
Multiple SARS-CoV-2 introductions into the Gundersen Health System service area (SW Wisconsin, NE Iowa, SE Minnesota). Viruses sequenced in this study are indicated in colored clades in this radial time-resolved phylogenetic tree. Reference sequences from other laboratories from elsewhere in MN, WI, IA and IL are colored gray. With the exception of the Postville cluster, clusters are named for the county in which the first local case was sequenced.

Notably, the majority of viral introductions did not give rise to outbreaks consisting of sustained transmission chains. Often, where multiple cases are clustered on the tree, they consist of individuals sharing the same household. A striking exception was a cluster of cases detected in Allamakee County, Iowa (Fig 1, lower right). This cluster is associated with multiple individuals associated with a meatpacking plant in the small town of Postville, Iowa (estimated population 2,083).

Since 4/6/2020 we have sequenced 27 viral cases that are attributed to this substrain (typically identified by the SARS-CoV-2 polymorphisms C9866T, A14747G and A20755C). Allamakee County reported 119 cases between late March and 5/26/2020, so we believe that the sequences reported here represent only a sample of the cases arising from this outbreak. In addition, factors common in the meatpacking workforce in this region (non-English speaking, under- or uninsured, low socio-economic status, undocumented immigration status) may also adversely affect the ability to achieve a comprehensive case count.

To estimate the origin of this viral substrain, we examined the position of this cluster relative to other sequenced specimens from other regions on NextStrain.org (Fig 2). To facilitate understanding of the origin and spread of these viral substrains, data are resolved by either time (diagnosis date, Fig 2A) and by divergence (number of non-reference mutations, Fig 2B). This allowed us to ascertain the order of the acquisition of these non-reference variants (first A20755C, then A14747G, followed by C9866T) and the divergence that occurred among sequenced substrains over time. The substrain ancestral to most of these viruses (containing only A20755C) was first detected in specimens obtained in Connecticut (CT-Yale) and in Israel in mid-March (Fig 2A). Analysis of divergence in this portion of the tree (Fig 2B) shows 8 identical viral sequences (5 from Israel, 1 from Russia, 1 from Connecticut and 1 from Maryland; These specimens are aligned vertically in Fig 2B). Additional slightly more divergent sequences within this group were from similar locations (New York, Connecticut, Israel). These data implicate the origin of the Postville virus being in the northeastern United States with some overlap with Israel. The outgroup to the sequences shown in this figure is mostly comprised of specimens sequenced in the USA so it seems most likely that US-to-Israel rather than Israel-to-US spread contributed to the observed close association of these specimens on this tree. After the acquisition of the A14747G mutation, the tree splits in two, with one variant defining the Postville cluster (C9866T) and another variant (C16658T) defining a cluster of cases identified in Arizona. The inferred ancestry of this Postville cluster in the northeastern USA in early March is consistent with media reports of initial COVID19-positivity in three family members associated with the meatpacking plant who returned to Postville from New York City in early March (9).

**Figure 2.**
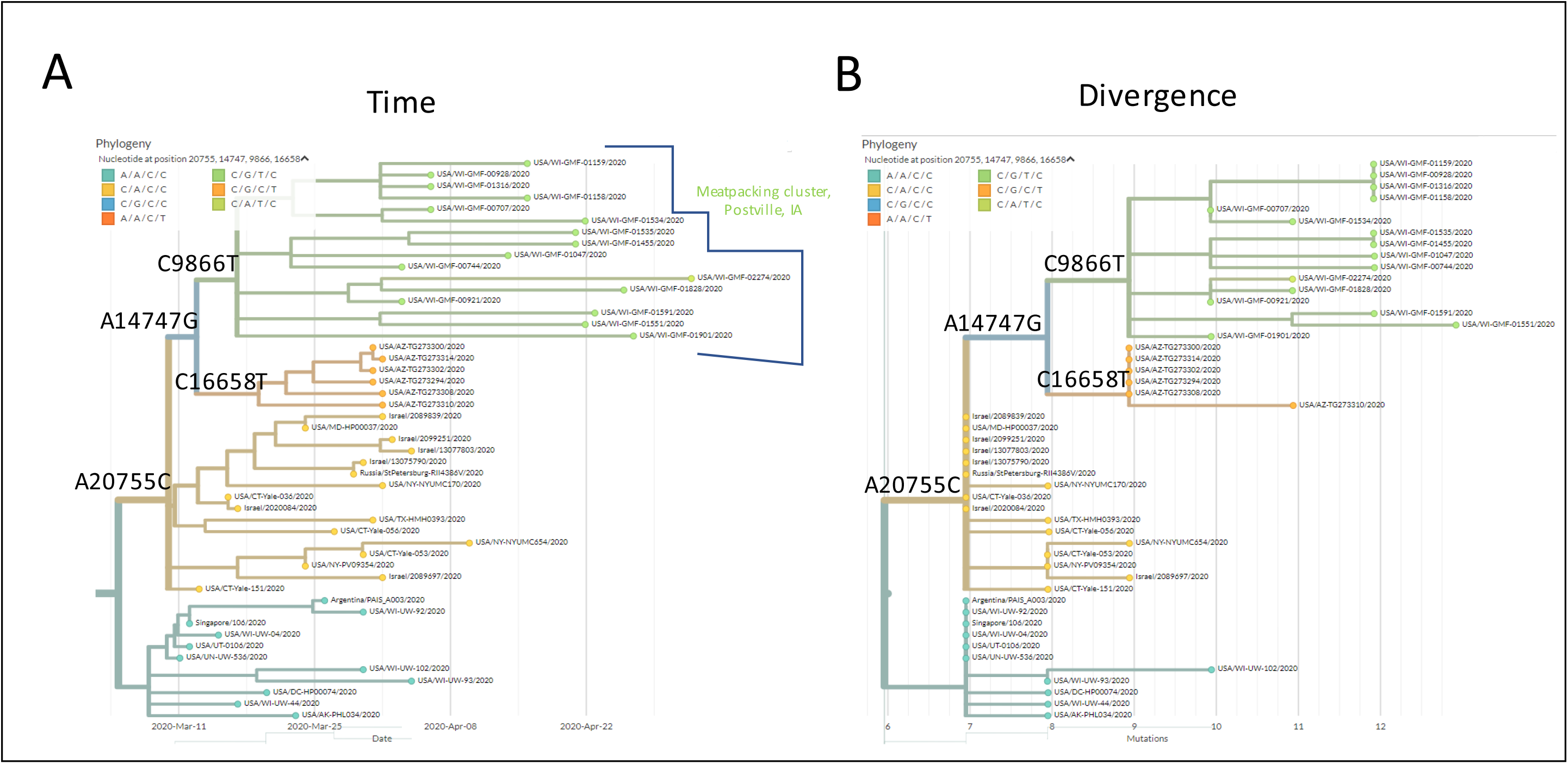
Phylogenetic trees showing the relationship between the Postville, IA cluster (green) and SARS-CoV-2 sequences from elsewhere. The trees are either time resolved (date of diagnosis on X-axis, Fig 2A) or divergence-resolved (number of new mutations on X-axis, Fig 2B).

In the early phase of this outbreak (first two weeks), the majority of cases sequenced from this substrain were among plant workers or individuals living in their households. The first 16 cases were from Allamakee County (13 cases) or neighboring Winneshiek County (3 cases). Although public health departments typically do not release zip code level details, data published in the media on 5/12/2020 indicated that Postville accounted for the overwhelming majority of cases (87 out of 97) in Allamakee county at that time (10). Thereafter, numerous cases were detected by sequencing in nearby and, later, more distant, communities among individuals with no known direct link to the meatpacking workforce (See Table 1 and Fig 3). In mid-May, we identified a case linked to this cluster in Trempealeau County, WI which is more than 100 miles distant from the Postville epicenter. Cases traceable to this outbreak have now been detected in seven counties in three states, affecting a total of 13 municipalities. This region spans 185 square miles.

**Table 1.** Counties and cases associated with the Postville, Allamakee County, Iowa outbreak.

| State | County | Number of cases sequenced in this study | Number of municipalities | Total cases reported by county (at 5/26/20) |
| --- | --- | --- | --- | --- |
| Iowa | Allamakee | 15 | 3 | 119 |
| Iowa | Winneshiek | 3 | 3 | 23 |
| Wisconsin | Vernon | 4 | 2 | 17 |
| Wisconsin | Grant | 1 | 1 | 87 |
| Wisconsin | Crawford | 2 | 2 | 26 |
| Wisconsin | Trempealeau | 1 | 1 | 21 |
| Minnesota | Fillmore | 1 | 1 | 17 |

**Fig 3.**
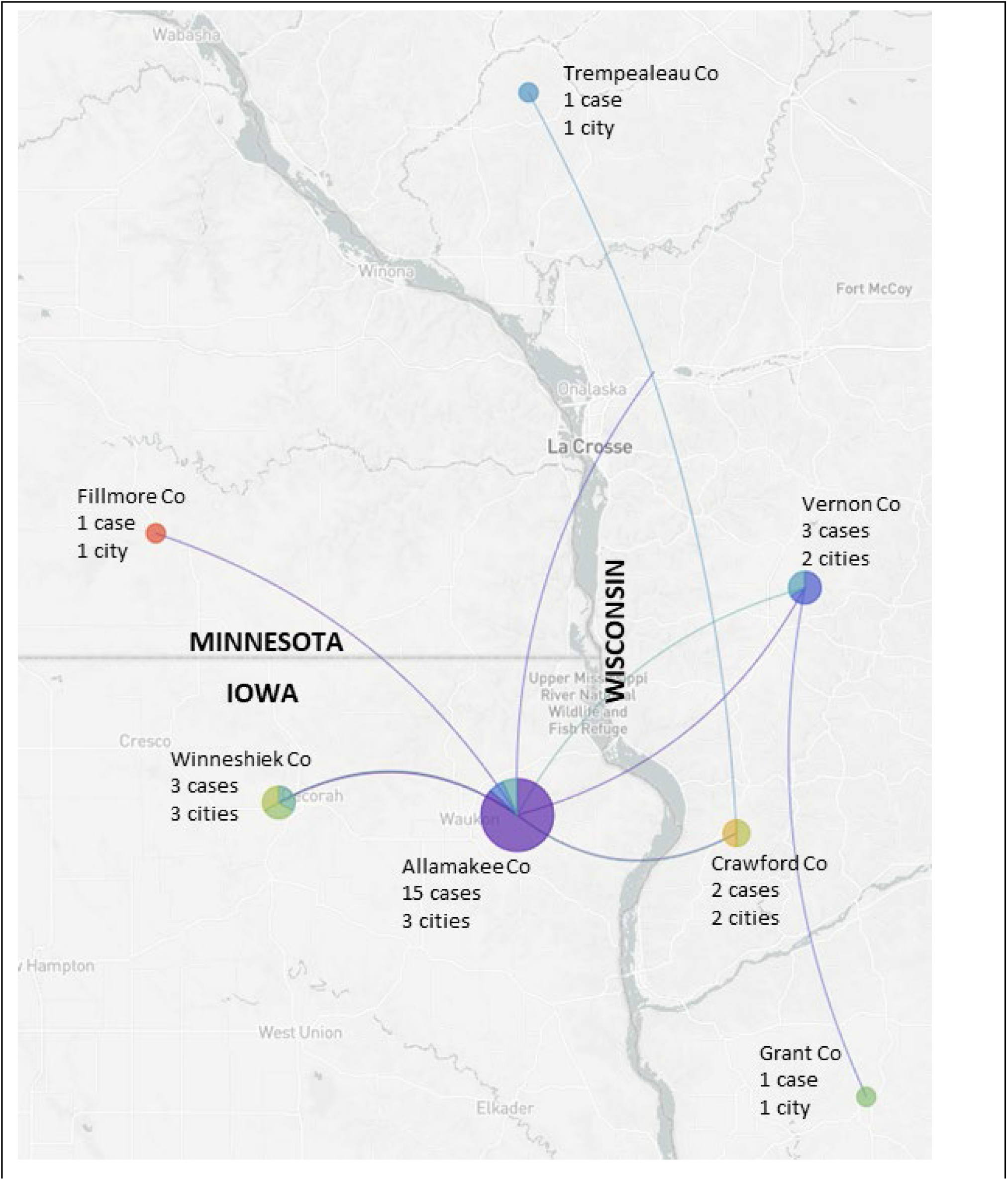
Interregional spread of SARS-CoV-2 from the Postville, IA (Allamakee County) outbreak. The largest cluster is in Postville itself. Lines between municipalities represent possible transmission chains imputed by NextStrain. Data are presented at the county level (charts geolocated at the center of each county), with the number of affected cities listed in each case. Pie charts indicate the proportion of cases for that county that are located in the affected cities.

Although documenting the spread of this viral substrain was the primary focus of the study, we performed an exploratory analysis of demographic and outcome data on affected individuals, summarized in Table 2. The small sample size limited the statistical power of the analysis. We stratified data between the outbreak county (Allamakee county; 15 cases) versus areas of subsequent spread (other counties; 12 cases). We noted a non-significant trend toward older age in counties beyond the epicenter. More individuals in the latter counties had significant relevant co-morbidities (diabetes and/or chronic kidney disease, p = 0.0085). The hospital admission rate in the other counties was also numerically higher (58.3% v 26.7%, p = ns). One individual died from COVID19.

**Table 2.** Characteristics of cases, stratified by epicenter county (Allamakee) v others.

|  |  | All Individuals (n = 27) | Allamakee County (n = 15) | Other counties (n = 12) | p-value (Allamakee v others) |
| --- | --- | --- | --- | --- | --- |
| **Age** | Minimum | 7 | 20 | 7 |  |
|  | Median | 40 | 38 | 56.5 | 0.16 |
|  | Maximum | 80 | 64 | 80 |  |
| **Sex** | Male | 13 (48.1%) | 5 (33.3%) | 8 (66.7%) | 0.12 |
|  | Female | 14 (51.9%) | 10 (66.7%) | 4 (33.3%) |  |
| **Comorbid conditions** | Present | 8 (29.6%) | 1 (6.7%) | 7 (58.3%) | 0.0085 |
| **(Diabetes or CKD)** | Absent | 19 (70.4%) | 14 (93.3%) | 5 (41.7%) |  |
| **Hospital Admission** | Yes | 11 (40.7%) | 4 (26.7%) | 7 (58.3%) | 0.13 |
|  | No | 16 (59.3%) | 11 (73.3%) | 5 (41.7%) |  |

The extent to which viral substrain differences may result in different clinical symptoms has been the subject of speculation. In this series of 27 patients with clearly confirmed infection with a single substrain, the patients had the following symptoms at presentation documented in their medical records: Fever (59.3%), cough (51.9%), myalgia (29.6%), dyspnea (22.2%), diarrhea (22.2%), fatigue (18.5%), anosmia (18.5%), headache (18.5%) and sore throat (18.5%), indicating substantial initial symptom variability among individuals infected with essentially identical viruses. This variability, particularly the high proportion of individuals lacking fever, highlights the challenges inherent in deploying effective workforce symptom screening.

## DISCUSSION

Contemporaneous with the outbreak reported here, several outbreaks at meat processing plants have been reported across the United States (11). While the importance of meat to the national food supply has been emphasized at the highest levels of government, the full implications of such policies require assessment of impacts not just to individuals designated “essential workers” but to their families, communities and broader regional populations.

The genomic epidemiology in the present study documents not just the effects of unmitigated spread of SARS-CoV-2 within a meatpacking plant, but the collateral damage resulting from widespread dissemination of this disease from a meat-packing epicenter across a large midwestern region. Unlike most of the SARS-CoV-2 introductions to our region which resulted in minimal spread (Fig 1), this particular introduction grew explosively. Review of the clinical histories of individuals tested in this study revealed anecdotes of numerous additional household members exhibiting symptoms of COVID19 but who were not tested, underlining the fact that the cases presented here represent only a subset of the cases likely attributable to this source.

The ability to move beyond simple identification of positive cases to unambiguously identifying patterns of case-to-case spread and cluster formation will improve the ability of the healthcare system (both hospitals and public health departments) to understand and monitor the emergence and spread of SARS-CoV-2 infections from new clusters as they are seeded. This may allow discovery of weak points in current disease control strategies in ways that might not otherwise be possible (e.g. linking of meatpacking cluster with a new case more than 100 miles distant). Such weak points may be amenable to mitigation by specific additional social, structural, or technical interventions.

The public health infrastructure in the United States has suffered from systematic underinvestment for many years (12). A comprehensive and sustained effort to track and mitigate SARS-CoV-2 spread may be limited by capacity and resource issues, which may be particularly acute in rural counties, but also by the ability of public health officials to communicate efficiently across county and state lines. A significant strength of the current study is the co-location of the sequencing team with the molecular diagnostics laboratory which was among the earliest laboratories to provide SARS-CoV-2 testing across this wide region. By sequencing viral genomes from a region spanning 21 counties in 3 states, our team has been able to achieve an overview of interregional spread that would be most challenging for smaller entities. Dissemination of such information to local county health departments may assist them in more efficiently targeting limited resources to high yield interventions to mitigate SARS-CoV-2 spread.

## CONCLUSIONS

Overall, these data, which almost certainly undersample the true extent of disease spread, document the regional consequences of early failures to mitigate SARS-CoV-2 contagion in an industrial setting.

## Data Availability

Consensus sequence data on all viral substrains has been submitted to GISAID.

https://www.gisaid.org/

## ACKNOWLEDGEMENTS

This work was supported by the Gundersen Medical Foundation. PK holds the Dr. Jon & Betty Kabara Endowed Chair in Precision Oncology. We thank the members of the Gundersen Medical Foundation’s Molecular Diagnostics Laboratory for providing the specimens used in this study.

## REFERENCES

1. Zhu N, Zhang D, Wang W, Li X, Yang B, Song J, et al. A Novel Coronavirus from Patients with Pneumonia in China, 2019. N Engl J Med 2020;382:727–33

2. Wang C, Liu Z, Chen Z, Huang X, Xu M, He T, et al. The establishment of reference sequence for SARS-CoV-2 and variation analysis. J Med Virol 2020

3. van Dorp L, Acman M, Richard D, Shaw LP, Ford CE, Ormond L, et al. Emergence of genomic diversity and recurrent mutations in SARS-CoV-2. Infect Genet Evol 2020;83:104351

4. Li H, Handsaker B, Wysoker A, Fennell T, Ruan J, Homer N, et al. The Sequence Alignment/Map format and SAMtools. Bioinformatics 2009;25:2078–9

5. Kim D, Paggi JM, Park C, Bennett C, Salzberg SL. Graph-based genome alignment and genotyping with HISAT2 and HISAT-genotype. Nat Biotechnol 2019;37:907–15

6. Cavener DR. Comparison of the consensus sequence flanking translational start sites in Drosophila and vertebrates. Nucleic Acids Res 1987;15:1353–61

7. Shu Y, McCauley J. GISAID: Global initiative on sharing all influenza data - from vision to reality. Euro Surveill 2017;22

8. Hadfield J, Megill C, Bell SM, Huddleston J, Potter B, Callender C, et al. Nextstrain: real-time tracking of pathogen evolution. Bioinformatics 2018;34:4121–3

9. TheYeshivaWorld. 2020 AGRI-RESPONDS: Members Of Postville Frum Community Diagnosed With Coronavirus. < https://www.theyeshivaworld.com/news/headlines-breaking-stories/1840313/panic-in-postville-iowa-3-members-of-frum-community-diagnosed-with-coronavirus.html>.

10. Murray I. 2020 Postville Has Vast Majority Of Allamakee’s COVID Cases, Councilman Finds.<https://iowastartingline.com/2020/05/12/postville-has-vast-majority-of-allamakees-covid-cases-councilman-finds/>.

11. Dyal JW, Grant MP, Broadwater K, Bjork A, Waltenburg MA, Gibbins JD, et al. COVID-19 Among Workers in Meat and Poultry Processing Facilities - 19 States, April 2020. MMWR Morb Mortal Wkly Rep 2020;69

12. Maani N, Galea S. COVID-19 and Underinvestment in the Public Health Infrastructure of the United States. Milbank Q 2020

